# SPECIFIC DETECTION OF SARS-COV-2 VARIANTS B.1.1.7 (ALPHA) AND B.1.617.2 (DELTA) USING A ONE-STEP QUANTITATIVE PCR ASSAY

**DOI:** 10.1101/2021.10.11.21264831

**Authors:** Oran Erster, Ella Mendelson, Areej Kabat, Virginia Levy, Batya Menasheh, Hadar Asraf, Roberto Azar, Yaniv Ali, Efrat Bucris, Dana Bar-Ilan, Orna Mor, Michal Elul, Michal Mandelboim, Danit Sofer, Shai Fleishon, Neta S Zuckerman, Itay Bar-Or

**Affiliations:** Central Virology Laboratory, Public Health Services, Ministry of Health, Chaim Sheba Medical Center, Ramat Gan, Israel; School of Public Health, Sackler Faculty of Medicine, Tel-Aviv University, Tel-Aviv, Israel

**Author notes:** These authors contributed equally to this study (Neta S Zuckerman, Itay Bar-Or).

**Keywords:** SARS-COV-2, RT-qPCR, Alpha (B.1.1.7), Delta (B.1.617.2), classification

## Abstract

In this report, we describe the development of an RT-qPCR assay, termed Alpha Delta assay, which can detect SARS-COV-2 (SC-2) and distinguish between the Alpha (B.1.1.7) and Delta (B.1.617.2) variants. The Alpha- and Delta-specific reactions in the assay target mutations that are strongly linked to the target variant. The Alpha reaction targets the D3L substitution in N gene, and the Delta reaction targets the spike gene 156-158 mutations. Additionally, we developed a second Delta-specific assay, used as a confirmatory test for the Alpha Delta assay that targets the 119-120 deletion in the Orf8 gene. Both reactions have similar sensitivities of 15-25 copies per reaction, similar to the sensitivity of commercial SC-2 detection tests. The Alpha Delta assay and the Orf8-119del assay were successfully used to classify clinical samples that were subsequently analyzed by whole genome sequencing. Lastly, we show that the Alpha Delta and Orf8-119del assays correctly identified the presence of Alpha and Delta lineages RNA in wastewater samples. This study provides a rapid, sensitive and cost-effective tool for detecting and classifying two worldwide dominant SC-2 variants. It also highlights the importance of a timely diagnostic response to the emergence of new SC-2 variants with significant consequences on global health.

## 1. INTRODUCTION

The emergence of new SC-2 variants has been a major concern due to their potential to increase morbidity and mortality, thereby causing further difficulty in fighting and containing the worldwide COVID19 pandemic [1,2,3]. Since the end of 2020, more than 20 SC-2 variants with distinct genomic signature have been identified worldwide, some of these variants divided into additional subclades (https://covariants.org/, https://cov-lineages.org/lineage_list.html). Some of them were characterized by markedly increased infectivity, which led to recurring outbreaks and “take-over” effect of specific variants, such as B.1.1.7 (Alpha) in a matter of 4-5 weeks [1]. Other variants, such as B.1.351 (Beta) and P1 (Gamma) spread rapidly and showed a significant potential to become dominant [4], but were eventually pushed aside by others. Variant B.1.617, initially identified in India on late 2020, spread rapidly and one of its sub-lineages-B.1.617-2 became a globally dominant variant within a few months. The transmission pattern of Alpha and Delta variants suggests that they both spread from their country of origin, rapidly reaching North and South America, Europe, Africa and Australia [4]. In Israel, two large morbidity waves were associated with the introduction of new variants. First, the Alpha variant high morbidity wave, which started in mid-December 2020 and declined by mid-March 2021 following the successful nation-wide immunization campaign with two doses of the BNT162-b2 vaccine [5]. Then, the current high morbidity wave dominated by the Delta variant, which started in July 2021 in parallel to vaccines waning immunity, and continues [6]. Immunological studies demonstrated that some of the emerging variants have the potential to evade, at least partially, the neutralizing effect of both convalescent patients and vaccinated individuals [2,3,7,8,9]. The spreading of such variants my therefore compromise the effectiveness of global vaccination efforts, aimed to contain the COVID19 pandemic.

A fundamental pillar of the campaign against COVID19 is the ability to detect the presence and identity of SC-2 RNA in both infected individuals and environmental samples. This pressing need led to an unprecedented number of commercial detection kits based on molecular and serological principles, with varying degrees of sensitivity, throughput, reliability and turnaround time [10, 11,12 and references therein]. Due to their epidemiological and medical importance, the rapid identification of circulating variants becomes a central tool in the fight against the pandemic. In addition to diagnosis of clinical samples, the monitoring of SC-2 RNA in environmental samples, mostly, but not exclusively, wastewater, proved as an important tool [6,13,14]. Detection of SC-2 RNA in such samples is exceptionally challenging due to their very low abundance and the presence of PCR inhibitors, which render this approach even more difficult. Several reports describe the successful identification of SC-2 RNA in environmental samples, and some describe the classification of the examined samples, using sequencing [6,15,16]. Classification of samples using sequencing requires sufficient quality and quantity, which are often not available, with environmental samples. Moreover, this approach is currently much more expensive compared with PCR-based assays, and cannot be scaled-up readily, to facilitate rapid screening of a large number of samples efficiently.

In order to address the need for rapid classification of SC-2 positive samples, several manufacturers released sets of molecular tests that specifically detect key mutations that are associated with increased transmissibility or vaccine resistance (www.seegene.com, www.thermofisher.com, www.kogene.co.kr). However, most of these mutations are common to multiple variants and therefore cannot be used to assign a sample of interest to a certain lineage with high degree of confidence using a single test. We recently developed quantitative PCR (qPCR)-based molecular tests that target mutations that are strongly associated with variants Alpha and Beta, thereby enabling rapid classification of such samples, with high confidence. We demonstrated that the selected target mutations successfully reflect the lineage of the examined sample, as confirmed by Sanger and whole genome sequencing [17]. In this report, we describe the development and utilization of two qPCR reactions that target mutations that are, thus far, unique to variant B.1.617 (Delta lineage). We combined one of these reactions with the N_D3L_ reaction that detects the Alpha variant, and the inclusive E-sarbeco reaction [18] we described previously, to a new selective multiplex assay. This assay can determine if the examined samples is of lineage Alpha, Delta, or none of them. We show that the new multiplex assay, which we term “Alpha Delta assay”, is rapid, sensitive and reliable, as confirmed by sequencing. Finally, we demonstrate the capability of the new assay to detect and classify SC-2 RNA extracted from environmental samples.

## 2. MATERIALS AND METHODS

### 2.1. Preparation of clinical and environmental samples

Clinical samples were prepared during the laboratory diagnostic routine from nasopharyngeal swabs, as described before [17]. RNA was extracted using either MagNA-Pure 96 system (Roche, https://lifescience.roche.com/) or PPS MagLEAD system (http://www.pss.co.jp/english/). The extractions were used for routine clinical diagnostics and were incubated at -80°C for long-term storage prior to the novel assay development. Environmental samples were processed as described before [6]. Briefly, sewage samples were centrifuged and filtered before extraction was commenced with either Nuclisense EasyMag (Biomerieux, https://www.biomerieux-usa.com/clinical/) or EMag extraction systems. Immediately after extraction, all samples were preserved at -80°C until PCR testing was performed.

### 2.2. Cell culture

In order to culture positive SC-2 samples, the medium from the selected collection tubes was filtered using a 0.22µM filter (https://www.merckmillipore.com/) and used for inoculation of Vero E6 cells. The cells were allowed to reach 70% confluence and were then incubated for 1 hour at 37°C with 300µl of the filtered medium. After 1 hour, the medium was removed and the cells were incubated in MEM-EAGLE medium with 2% FCS. Cells were monitored daily for the presence of cytopathic effect (CPE). Upon CPE onset, the culture medium was collected and viral RNA was extracted. Resulting RNA extractions were kept at -80°C until use.

### 2.3. Design of B.1.671-specific RT-qPCR assays

Sequences classified as B.1.617 were obtained from the GISAID database (https://www.epicov.org/epi3/frontend#2d1c1e) and were analyzed by alignment with reference sequence NC_045512 to identify mutated regions that can be used for differential analysis. Uniqueness of the selected mutations was confirmed by performing global analysis for the selected mutations in the NextStrain website, to identify other lineages with identical mutations (https://nextstrain.org/ncov/gisaid/global). Corresponding primers and probes that detect only the mutated sequences were designed for each region, and examined *In silico* for secondary structure formation, specificity and compatibility with qPCR assay using the Geneious software (https://www.geneious.com/) and using the NCBI BLAST (https://blast.ncbi.nlm.nih.gov/Blast.cgi). The primers and probes used in this study are detailed in **Table 1**.

**Table 1.**
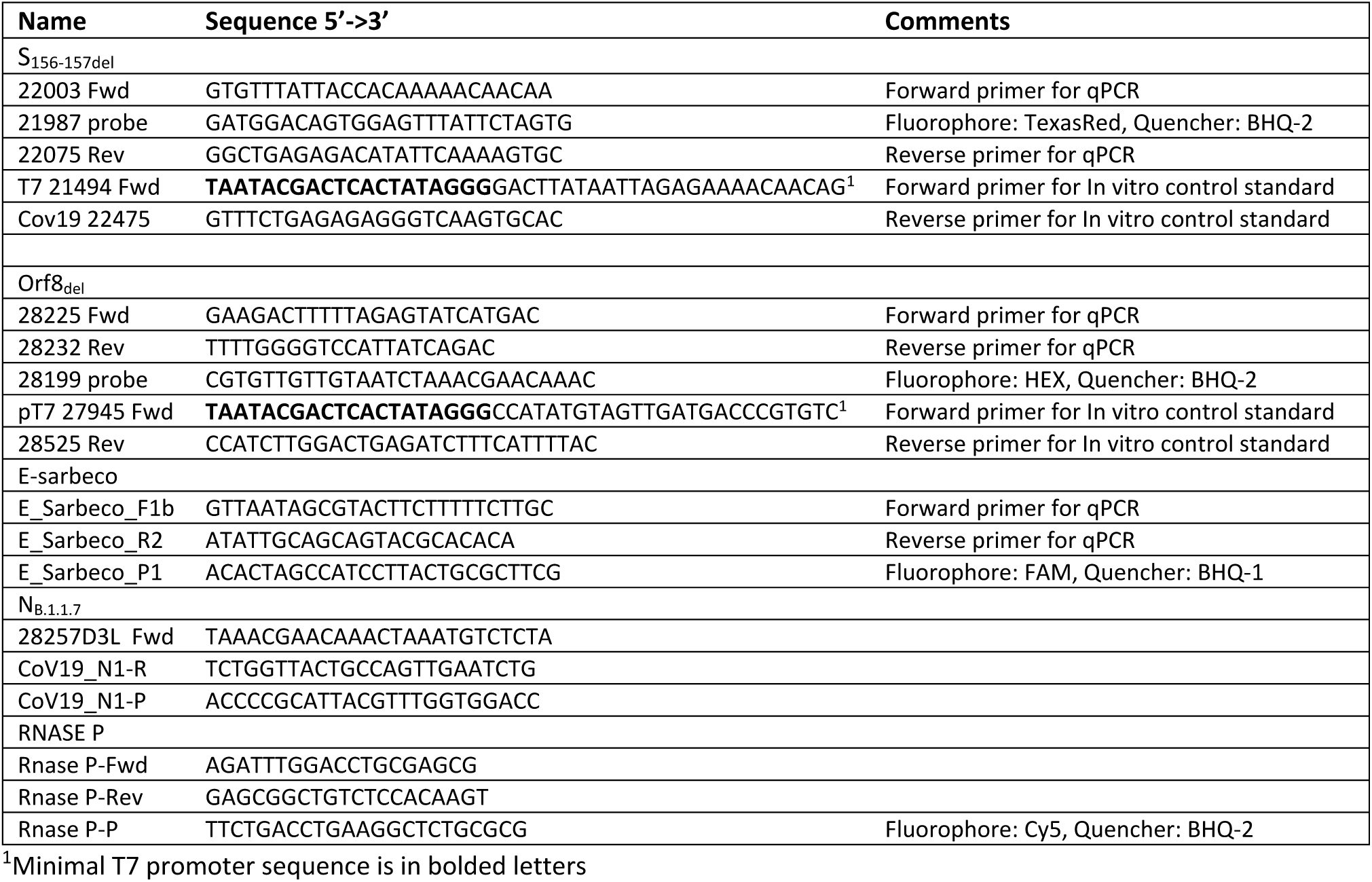
Primers and probes used for the development of the S_156-157del_ and Orf8_119-120del_ assays

### 2.4. RT-qPCR assay

MDX106 Inhibitor-tolerant RT-qPCR mix was from Bioline (Bioline, https://www.bioline.com/). Primers and probes were either from Metabion (https://www.metabion.com/) or from Merk-Sigma Israel (https://www.sigmaaldrich.com/IL/en). Reaction components were assembled as described in **Table 2**, using the MDX106 Inhibitor-tolerant mix. The reactions were run in CFX-96 thermal cycler (Bio-Rad, https://www.bio-rad.com/), using the following profile:

**Table 2.** Reaction mix composition for the S_157del_ and Orf8_119del_ assays.

| **B617 S<sub>156-157del</sub> +E + RNSP** | **Conc / Rxn** | **Volume/reaction (μl)** |
| --- | --- | --- |
| Meridian MDX 4X mix | X1 | 6.25 |
| H<sub>2</sub>O |  | 8.55 |
| 28225 Fwd | 500nM | 0.25 |
| 28232 Rev 40uM | 500nM | 0.25 |
| 28199 Probe 20uM | 250nM | 0.25 |
| E-Sarbeco-F1b 40μM | 400nM | 0.25 |
| E-Sarbeco-R 40μM | 400nM | 0.25 |
| E-Sarbeco-P FAM 20μM | 200nM | 0.25 |
| RNasP-F | 2X ( of 300nM) | 0.25 |
| RNasP-R | 2X ( of 300nM) | 0.25 |
| RNasP-P / Cy-5 | 2X ( of 200nM) | 0.2 |
| Total |  | 16.3 |

| **B617 Orf8<sub>119-120del</sub> multiplex mix** | **Final concentration** | **Volume/reaction (μl)** |
| --- | --- | --- |
| Meridian MDX 4X mix | X1 | 6.25 |
| H<sub>2</sub>O |  | 9.25 |
| 28225 Fwd | 500nM | 0.25 |
| 28232 Rev 40uM | 500nM | 0.25 |
| 28199 Probe HEX 20uM | 250nM | 0.25 |
| E-Sarbeco-F1b 40μM | 400nM | 0.25 |
| E-Sarbeco-R 40μM | 400nM | 0.25 |
| E-Sarbeco-P FAM 20μM | 200nM | 0.25 |

45°C for 10’10”, 45X[95°C for 2’20”, 95°C for 4”, 60°C for 22”]. Fluorescence was recorded at 60°C, at the end of each amplification cycle. Results were analyzed using the CFX Maestro software (Bio-Rad).

### 2.5. Sanger sequencing of culture-derived and clinical samples

The N-terminal domain (NTD) of the SC-2 Spike gene and the C-terminal domain (CTD) of SC-2 Orf8 were amplified using the primer pairs detailed in **Table 1**. Primer pair T7-21494 Fwd + Cov19 22475 were used for the Spike region amplification, and primer pair pT7 27945 Fwd + 28525 Rev were used for the Orf8 region amplification. PCR was performed using the PCRBIO 1-Step Go RT-PCR Kit (https://pcrbio.com/row/products/pcr/pcrbio-1-step-go-rt-pcr-kit/) according to the manufacturer’s instructions. The PCR products were resolved in 1.2% agarose gel electrophoresis to confirm adequate amplification. Each product was then used as template for dye labeling reaction using the BigDye kit (https://www.thermofisher.com/) according to the manufacturer’s instructions. Sequencing was performed using the ABI 3500 genetic analyzer (https://www.thermofisher.com/).

### 2.6. Generation of In-vitro transcribed standard RNA

RNA standards of the reaction targets were designed and synthesized as described before (Erster *et al*. 2021). The primers for the amplification of the S_157del_ and Orf8_119del_ reaction targets are detailed in **Table 1**. RT-PCR was performed with Fwd primers containing the minimal T7 promoter sequence. Resulting PCR products were purified using the Machery-Nagel NucleoSpin Gel and PCR Clean-up kit according to the manufacturer’s instructions (https://www.mn-net.com/). The purified products were then used for In vitro transcription with the T7-Megascript kit according to the manufacturer’s instructions (Thermo Fisher, https://www.thermofisher.com/). The concentration of the resulting RNA was measured using the NanoDrop spectrophotometer (https://www.thermofisher.com/il/en/). All reaction products were kept at -80°C until use.

### 2.7. Evaluation of analytical and clinical sensitivity of the Alpha-Delta assay

In order to facilitate rapid classification of the examined sample, four reactions were combined in a single assay. The following reactions were combined: inclusive E-sarbeco, which detects SC-2 RNA regardless of the variant (FAM fluorophore), N_D3L_ reaction, which detects the Alpha variant (HEX channel), S_157del_ reaction, which detects the Delta variant (TexasRed Channel), and an endogenous control reaction for the human RNAse P gene (Cy5 channel). The details of the E, N_D3L_ and RNAse P reactions were described previously (Erster *et al*. 2021). The combined multiplex was termed Alpha-Delta assay hereafter. The analytical sensitivity of the Alpha-Delta assay was evaluated using serial dilutions of the *In-vitro* transcribed targets of each reaction. All four targets were mixed together in an initial concentration of 10^−4^ ng/µl, and 10-fold dilutions were prepared in nuclease-free TE buffer pH7.5 (IDT, https://eu.idtdna.com/). The clinical sensitivity was evaluated by serially diluting positive samples in an extraction of a SC-2 – negative sample.

### 2.8. Collection and preparation of wastewater samples

Wastewater collection was based on composite automated sampler for 24 hour located in different wastewater treatment plants. The samples were transferred under cold conditions to the lab and were processed within 24 hours. Concentration process uses 20 ml of raw sewage in duplicates. The raw sewage was centrifuged at 4700g for 5 min. The supernatant was transferred to a new tube containing 0.5gr MgCl_2_ (0.26M). The tube was gently shaken for 5 min, followed by 0.45-μm pore-size and 47-mm diameter electronegative MCE membranes (Merck Millipore Ltd) filtration. The membrane was immediately transferred to a new tube containing 3 ml of lysis buffer (NucliSENS easyMAG). The tube was gently shaken to extract the trapped RNA virus. Total nucleic acids (NA) were extracted using the NucliSENS easyMAG system (bioM’rieux, Marcyl’Etoile, France) according to the manufacturer’s instructions. Extracted NA were eluted in 55 μl elution buffer and stored at −70 °C until sequencing.

### 2.9. Whole genome sequencing

COVID-seq kit was used for library preparation as per manufacturer’s instructions (Illumina, https://www.illumina.com). Library validation and mean fragment size was determined by Tapestation 4200 via DNA HS D1000 kit (Agilent, https://www.agilent.com/). Libraries were pooled, denatured and diluted to 10pM and sequenced on NovaSeq system (Illumina).

## 3. RESULTS

### 3.1. Design of the Spike_157del_ and Orf8_119del_ reactions

Alignment of genomes identified as the Delta lineage from the GISAID database (https://www.gisaid.org/) with SC-2 reference sequence NC_045512 showed that all Delta sequences contained a substitution in position deletion in positions 22045-22050, which translates into a.a. E to G substitution in position 156 and deletion in positions 157-158 in the Spike protein (**Figure 1A**). Accordingly, a specific reaction was designed to detect that deletion, denoted S_157del_ reaction hereafter, by using a probe that can only bind to the mutated sequence (21987 probe). In order to establish that the selected mutations were indeed unique to the Delta lineage, global analysis of SC-2 genomes was performed using the NextStrain website (https://nextstrain.org/), with the deletion in positions 22045-22050 as a search term. The resulting dendrogram showed that the double deletion was indeed unique to the Delta lineage and was absent from other known lineages (Supplementary **Figure 1A**).

**Figure 1.**
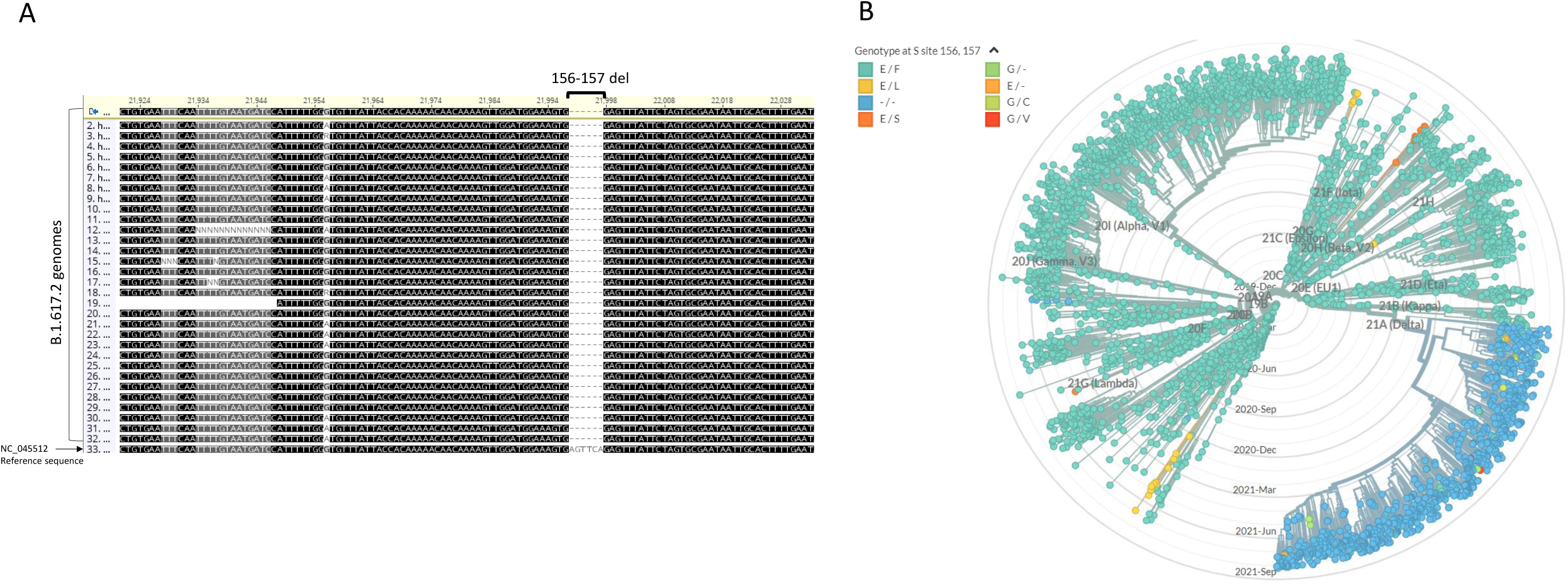
Idenfication of B.1.617-specific Spike gene deletion. (A) Alignment of 32 genomic sequences of lineage B.1.617 samples with reference sequence NC_045512 showed a deletion in nucleotide positions 22045-22050, corresponding to a.a. positions 156-157 in the protein sequence. The 156-157del gap is marked with brackets. (B) Global analysis of SC-2 genomes available in the NextStrain database (https://nextstrain.org/sars-cov-2/) showing that this 6-bp deletion is present only in the Delta lineage, highlighted in blue.

The Delta genomes alignment showed a second unique deletion, mapped to nucleotide positions 28248-28253 in reference sequence NC_045512C, at the C-terminal end of Orf8, spanning amino acids 119-120 of the protein (**Figure 2A**). Global genome analysis performed as described for the 22045-22050 deletion, showed that the deletion at positions 28248-28253 was also unique for Delta lineage (**Figure 2B**). A specific probe was designed to identify the deletion-containing sequence (28199). Secondary structure simulation suggested that the original probe sequence is likely to generate a strong stem-loop structure, thereby significantly reducing the reaction efficiency.in order to avoid this undesired outcome, a single G to T substitution was incorporated at position 10, in order to avoid the stem-loop formation (**Supplementary Figure S1**). Two primers were then designed to amplify the deletion region and complete the RT-qPCR assay. The sequences of the Orf8_119-120del_ primers and probe are detailed in **Table 1**. Sequencing of the C-terminal domain (CTD) of the Orf8 gene from the same four samples classified by the S_157del_ assay as Delta, confirmed the presence of the Orf8 119-120 deletion, which is the target of the Orf8_119del_ reaction (**Figure 2B**). Additional confirmation for the accuracy of the two reactions was performed by sequencing four samples that were identified as Delta-suspected. Each sample was sequenced in the S_157del_ region and the Orf8_119del_ region. As shown in **Supplementary Figure S2**, all sample contained both deletions.

**Figure 2.**
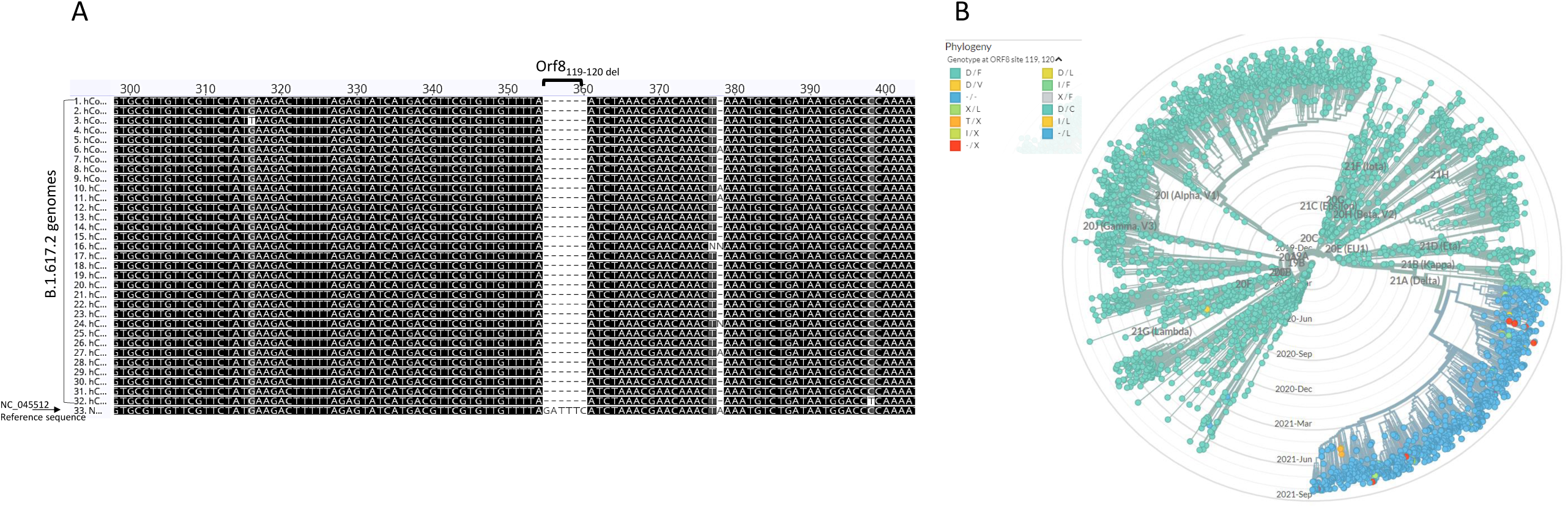
Idenfication of B.1.617-specific Orf8 gene deletion. (A) Alignment of 32 genomic sequences of lineage B.1.617 samples with reference sequence NC_045512 showed a deletion in nucleotide positions 28248-28253, corresponding to a.a. positions 119-120 at the C-terminal end of the protein sequence. The 119-120del gap is marked with brackets. (B) Global analysis of SC-2 genomes available in the NextStrain database (https://nextstrain.org/sars-cov-2/) showing that this 6-bp deletion is present only in the Delta lineage, highlighted in blue.

### 3.3. Development and evaluation of the Alpha-Delta assay

In order to facilitate detection of SC-2 RNA regardless of the lineage and identify the Alpha or Delta lineages in a single test, the three reactions were combined in one multiplex termed “Alpha-Delta assay”. A control reaction detecting human RNAseP gene was also included in the multiplex for an endogenous control, as described previously [17]. Analytical sensitivity of the Alpha-Delta assay was evaluated using serial dilutions of mixed RNA targets of the four reactions combined in the assay. Each dilution was examined in a triplicate. The limit of detection (LOD) for the SC-2 targets was determined to be 33.3 copies/reaction for the E-sarbeco reaction, 12.9 copies for the N_D3L_ reaction, and 14.5 copies for the S_157del_ reaction. The LOD for the hRNAse P reaction was 35.4 copies/reaction (**Supplementary Figure S3**). The Orf8_119del_ reaction was developed as a confirmatory assay for inconclusive Delta suspected samples. This reaction was used in combination with the E-sarbeco reaction that served as an inclusive SC-2 control reaction to ensure the presence of the SC-2 RNA. The analytical LOD of that reaction was determined to be 23.6 copies/reaction (**Supplementary figure S3**). The specificity of the Delta-specific reactions was tested using RNA extractions from lineages 19A/19B (Wuhan lineage), Alpha, Beta and Gamma. As detailed in Supplementary **Table S1**, all lineages were negative for both the S_157del_ and Orf8_119del_ reactions.

Analysis of the amplification curves of the different Delta assay variations showed that in some samples, mostly with a high RNA concentration (Cq≤22), a weak signal in the HEX channel was observed, assumingly resulting from low-affinity N_D3L_ reaction (**Figure 3D**). However, in these cases, the Cq values of the E-sarbeco reaction and the S_257del_ reaction (in Delta samples) were always at least 10 cycles earlier than that of the N_D3L_ reaction, thereby clearly indicating that the sample was not of the Alpha lineage. Both the S_157del_ and the Orf8_119del_ reactions were very specific and did not give a positive signal in non-Delta samples (**Figure 3**). In order to determine a possible mutual effect of the four reactions combined together, serial dilutions of Delta RNA from cultured cells were tested, showing similar sensitivity of both the E-sarbeco and S_157del_ reactions, thus confirming the maintained sensitivity of the Alpha-Delta multiplex (**Supplementary figure S4**).

**Figure 3.**
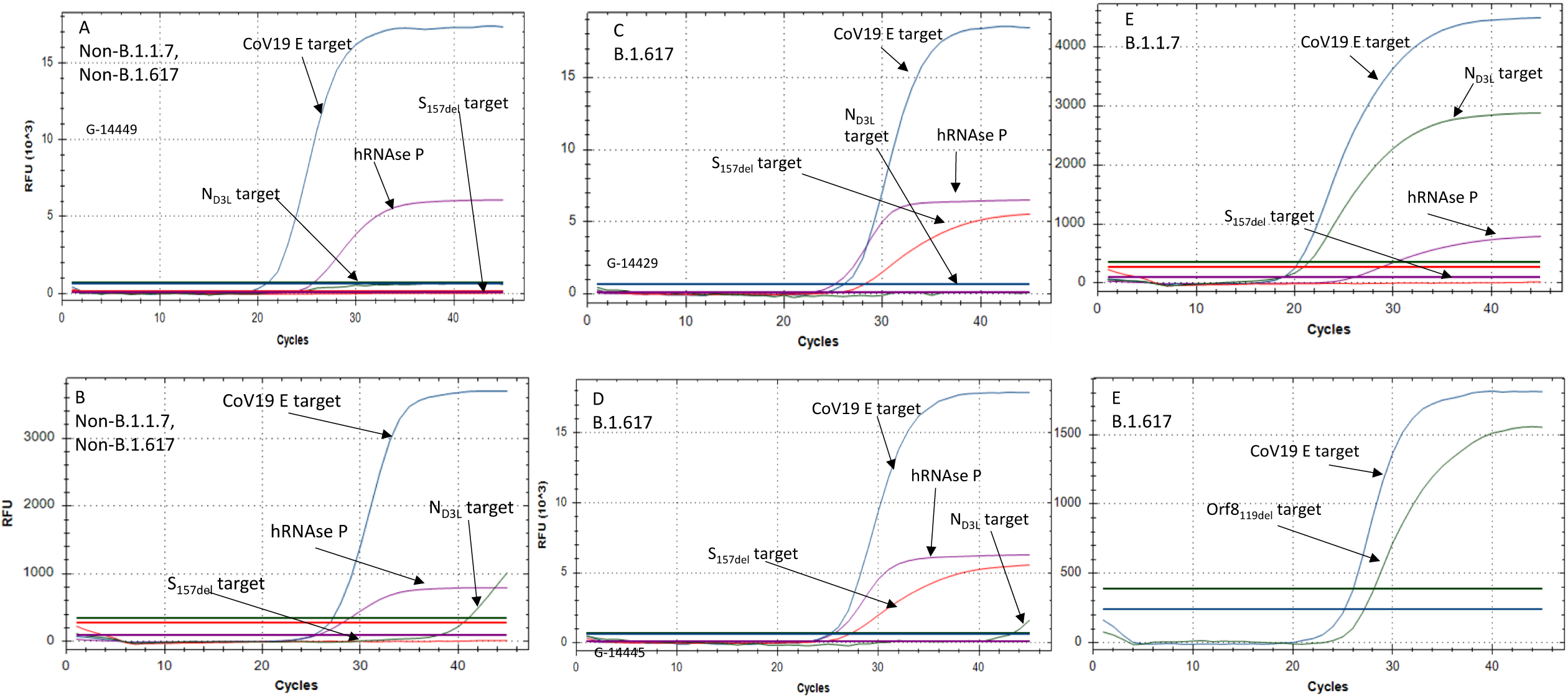
Amplification curves of the Alpha Delta and Orf8 assays. (A) Non-B.1.1.7, Non-B.1.617 sample with no background signal. (B) Non-B.1.1.7, Non-B.1.617 sample with ND3L reaction background. (C) B.1.617 sample with no background signal. (D) B.1.617 sample with ND3L reaction background. (E) B.1.617 sample reaction (F) B.1.617 E-sarbeco + Orf8199del reaction.

### 3.4 Examination of clinical samples using the Alpha-Delta assay followed by WGS analysis

Between January and May 2021, the prevalent lineage in Israel was Alpha. The Delta lineage began spreading in the country during mid-June 2021. The Alpha-Delta assay was therefore used to classify clinical samples whose lineage attribution was unknown, in order to enable rapid epidemiological investigations and to aid decision making regarding whole genome sequencing (WGS) prioritization. Later on, it was used as a routine test at the Israel Central Virology Laboratory. During June-August 2021, over 1,400 clinical samples were classified as Delta, Alpha or neither, using this assay. This classification was subsequently confirmed by Illumina WGS analysis. The details of 62 such samples selected randomly are listed in **Tables 3 and 4**. All samples classified by the Alpha-Delta qPCR assay as suspected Delta, were either identified as such by the WGS analysis, or classified as “no lineage” (**Table 3**). Those who were designated “no lineage” were mostly with Cq value higher than 32, and were all with 70% or less sequencing coverage **(Table 4**). These samples, which were classified as “Delta-suspected” by the Alpha-Delta assay, were positive for the Orf8 119-120 deletion, but the S_157del_ region was not fully sequenced. Comparison of the PCR results with the Illumina WGS analysis showed that of the 62 samples randomly selected, 19 were not classified by the WGS analysis, but were positive for the Delta-specific deletions, either the S157_del_ or the Orf8_119del_, or both (**Table 3**). Examination of the Cq values obtained for the WGS non-classified samples, showed that two samples had values of 27 and 29 (14957 and 14968, respectively), and all others were higher than 30 (**Table 3**). The sequence coverage of the unclassified samples ranged between 20.9% and 70% (**Table 4**). These data suggest that the positive identification of both the Orf8_119del_ and the S_157del_ mutations by the qPCR assay, enable a more definite classification of these samples, even in the absence of a complete sequencing coverage. The assay also detected a small number of Alpha lineage samples (two in the samples presented in **Tables 3, 4**), where the N_D3L_ mutation was detected by both the qPCR assay and the WGS analysis (**Tables 3, 4**).

**Table 3.** Cq values and classification of the Alpha-Delta assay, compared with the Pangolin classification (https://cov-lineages.org/resources/pangolin.html) as determined by the WGS analysis of 62 representative samples.

| Sample | CoV19 E | N<sub>D3L</sub> | S<sub>157del</sub> | ORF-8<sub>119del</sub> | Pangolin Clade | RT-PCR classification |
| --- | --- | --- | --- | --- | --- | --- |
| 14918 | 25 |  | 27 | 26 | B.1.617.2 | Suspected B.617.2 |
| 14920 | 36 |  | 38 | 40 | None | Suspected B.617.2 |
| 14921 | 26 |  | 28 | 26 | B.1.617.2 | Suspected B.617.2 |
| 14922 | 23 |  | 25 | 23 | B.1.617.2 | Suspected B.617.2 |
| 14923 | 17 |  | 19 | 18 | B.1.617.2 | Suspected B.617.2 |
| 14924 | 27 |  | 30 | 27 | B.1.617.2 | Suspected B.617.2 |
| 14925 | 36 |  | 37 | 34 | None | Suspected B.617.2 |
| 14926 | 22 |  | 24 | 22 | B.1.617.2 | Suspected B.617.2 |
| 14928 | 37 |  | N/A | 39 | None | Suspected B.617.2 |
| 14929 | 20 |  | 22 | 20 | B.1.617.2 | Suspected B.617.2 |
| 14931 | 31 |  | 33 | 30 | B.1.617.2 | Suspected B.617.2 |
| 14932 | 34 |  | 40 | 33 | None | Suspected B.617.2 |
| 14933 | 21 |  | 23 | 21 | B.1.617.2 | Suspected B.617.2 |
| 14934 | 29 |  | 31 | 29 | None | Suspected B.617.2 |
| 14936 | 35 |  | 40 | 33 | None | Suspected B.617.2 |
| 14937 | 21 |  | 23 | 21 | B.1.617.2 | Suspected B.617.2 |
| 14938 | 34 |  | 42 | 34 | None | Suspected B.617.2 |
| 14939 | 33 |  | 35 | 32 | None | Suspected B.617.2 |
| 14940 | 28 |  | 30 | 28 | B.1.617.2 | Suspected B.617.2 |
| 14941 | 24.68 | 25.51 | N/A | N/A | B.1.1.7 | Suspected B.1.1.7 |
| 14945 | 29 |  | 31 | 30 | None | Suspected B.617.2 |
| 14946 | 25 |  | 28 | 26 | B.1.617.2 | Suspected B.617.2 |
| 14947 | 25 |  | 27 | 26 | B.1.404 | Suspected B.617.2 |
| 14948 | 33 |  | 35 | 33 | None | Suspected B.617.2 |
| 14949 | 22 |  | 24 | 23 | B.1.617.2 | Suspected B.617.2 |
| 14950 | 27 |  | 29 | 28 | B.1.617.2 | Suspected B.617.2 |
| 14951 | 36 |  | 37 | 35 | None | Suspected B.617.2 |
| 14952 | 24 |  | 26 | 25 | B.1.617.2 | Suspected B.617.2 |
| 14953 | 22 |  | 24 | 22 | B.1.617.2 | Suspected B.617.2 |
| 14954 | 25 |  | 28 | 21 | B.1.617.2 | Suspected B.617.2 |
| 14955 | 25 |  | 27 | 26 | B.1.617.2 | Suspected B.617.2 |
| 14957 | 25 |  | 27 | 26 | None | Suspected B.617.2 |
| 14958 | 22 |  | 23 | 22 | B.1.617.2 | Suspected B.617.2 |
| 14960 | 27 |  | 29 | 27 | B.1.617.2 | Suspected B.617.2 |
| 14963 | 32 |  | 34 | 32 | None | Suspected B.617.2 |
| 14965 | 23 |  | 25 | 23 | B.1.617.2 | Suspected B.617.2 |
| 14966 | 23 |  | 15 | 23 | B.1.617.2 | Suspected B.617.2 |
| 14967 | 31 |  | 33 | 31 | None | Suspected B.617.2 |
| 14968 | 27 |  | 29 | 28 | None | Suspected B.617.2 |
| 14969 | 26 |  | 28 | 27 | B.1.617.2 | Suspected B.617.2 |
| 14970 | 25 |  | 28 | 26 | B.1.617.2 | Suspected B.617.2 |
| 14971 | 24 |  | 26 | 24 | B.1.617.2 | Suspected B.617.2 |
| 14972 | 32 |  | 39 | 32 | None | Suspected B.617.2 |
| 14973 | 28 |  | 29 | 28 | B.1.617.2 | Suspected B.617.2 |
| 14975 | 23 |  | 25 | 24 | B.1.617.2 | Suspected B.617.2 |
| 14978 | 21 |  | 21 | 21 | B.1.617.2 | Suspected B.617.2 |
| 14980 | 26 |  | 28 | 27 | B.1.617.2 | Suspected B.617.2 |
| 14999 | 27 | N/A | 29 | 29 | B.1.617.2 | Suspected B.617.2 |
| 15006 | 28 | NA | 30 | 29 | B.1.617.2 | Suspected B.617.2 |
| 15012 | 28 | NA | 30 | 29 | B.1.617.2 | Suspected B.617.2 |
| 15014 | 31 | NA | 32 | 33 | None | Suspected B.617.2 |
| 15019 | 25 | NA | 28 | 26 | B.1.617.2 | Suspected B.617.2 |
| 15024 | 31 | NA | 32 | 32 | None | Suspected B.617.2 |
| 15036 | 30 | NA | 31 | 31 | B.1.617.2 | Suspected B.617.2 |

**Table 3 - continued**
| **Sample** | **CoV19 E** | **N<sub>D3L</sub>** | **S<sub>157del</sub>** | **ORF-8<sub>119del</sub>** | **Pangolin Clade** | **RT-PCR classification** |
| --- | --- | --- | --- | --- | --- | --- |
| **15043** | 35 | NA | 36 | 36 | None | Suspected B.617.2 |
| **15046** | 27 | NA | 28 | 28 | B.1.617.2 | Suspected B.617.2 |
| **15048** | 29 | 28 | NA | NA | B.1.1 | Suspected B.1.1.7 |
| **15051** | 28 | NA | 30 | 29 | B.1.617.2 | Suspected B.617.2 |
| **15053** | 28 | NA | 29 | 28 | B.1.617.2 | Suspected B.617.2 |
| **15085** | 33 | NA | 33 | 32 | None | Suspected B.617.2 |

**Table 4.**
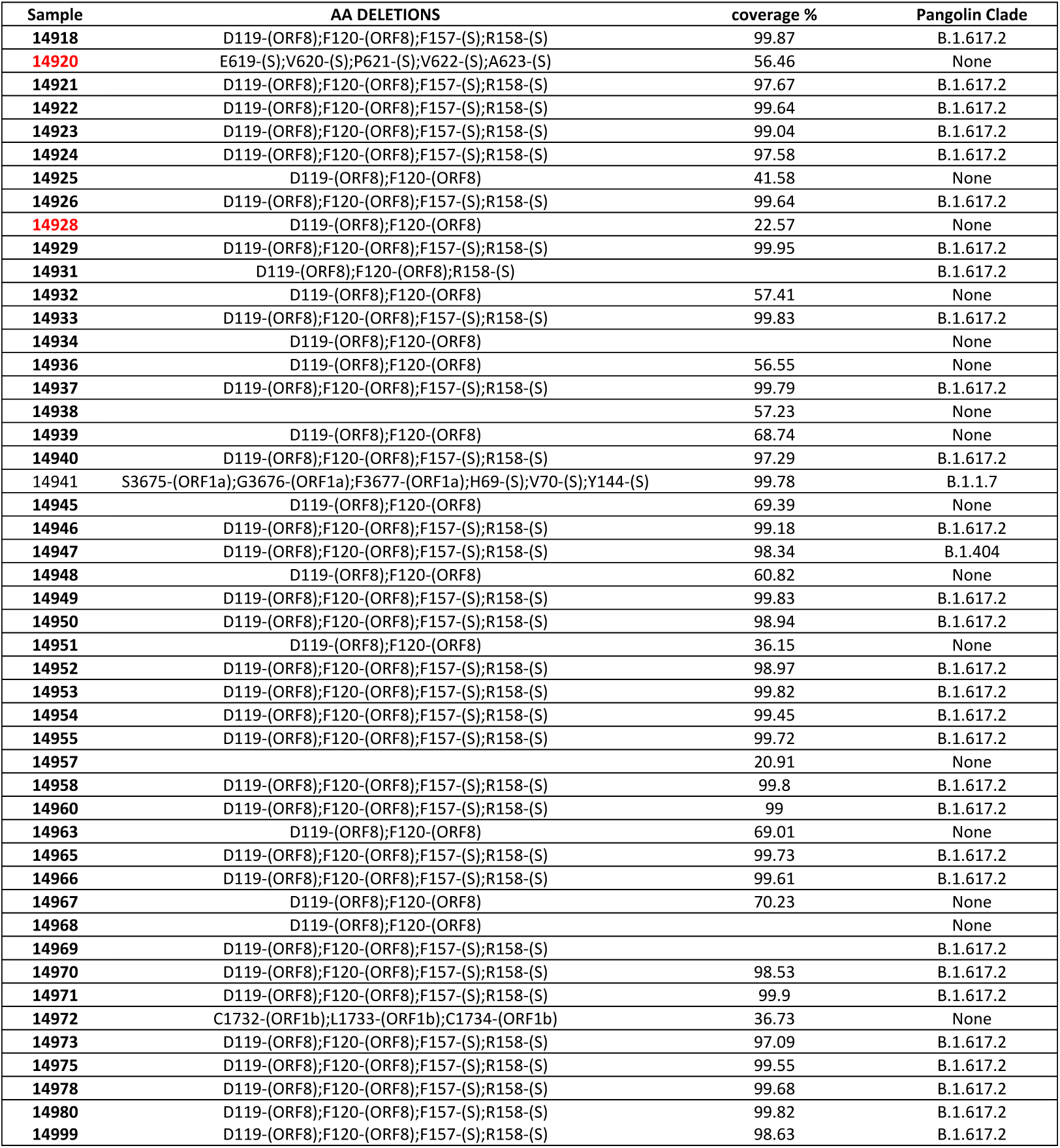
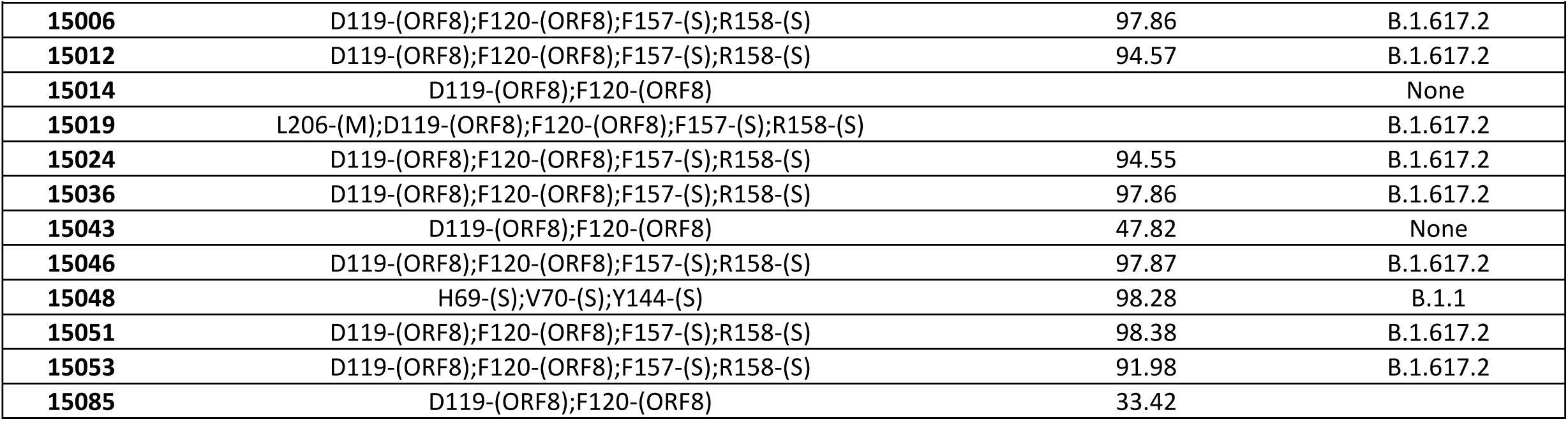
WGS Coverage, deletions sequenced by WGS analysis, and classification of clinical samples examined using the Alpha-Delta assay. The Alpha samples are highlighted. Delta NGS report 14918-14980 samples

| Sample | AA DELETIONS | coverage % | Pangolin Clade |
| --- | --- | --- | --- |
| 14918 | D119-(ORF8);F120-(ORF8);F157-(S);R158-(S) | 99.87 | B.1.617.2 |
| 14920 | E619-(S);V620-(S);P621-(S);V622-(S);A623-(S) | 56.46 | None |
| 14921 | D119-(ORF8);F120-(ORF8);F157-(S);R158-(S) | 97.67 | B.1.617.2 |
| 14922 | D119-(ORF8);F120-(ORF8);F157-(S);R158-(S) | 99.64 | B.1.617.2 |
| 14923 | D119-(ORF8);F120-(ORF8);F157-(S);R158-(S) | 99.04 | B.1.617.2 |
| 14924 | D119-(ORF8);F120-(ORF8);F157-(S);R158-(S) | 97.58 | B.1.617.2 |
| 14925 | D119-(ORF8);F120-(ORF8) | 41.58 | None |
| 14926 | D119-(ORF8);F120-(ORF8);F157-(S);R158-(S) | 99.64 | B.1.617.2 |
| 14928 | D119-(ORF8);F120-(ORF8) | 22.57 | None |
| 14929 | D119-(ORF8);F120-(ORF8);F157-(S);R158-(S) | 99.95 | B.1.617.2 |
| 14931 | D119-(ORF8);F120-(ORF8);R158-(S) |  | B.1.617.2 |
| 14932 | D119-(ORF8);F120-(ORF8) | 57.41 | None |
| 14933 | D119-(ORF8);F120-(ORF8);F157-(S);R158-(S) | 99.83 | B.1.617.2 |
| 14934 | D119-(ORF8);F120-(ORF8) |  | None |
| 14936 | D119-(ORF8);F120-(ORF8) | 56.55 | None |
| 14937 | D119-(ORF8);F120-(ORF8);F157-(S);R158-(S) | 99.79 | B.1.617.2 |
| 14938 |  | 57.23 | None |
| 14939 | D119-(ORF8);F120-(ORF8) | 68.74 | None |
| 14940 | D119-(ORF8);F120-(ORF8);F157-(S);R158-(S) | 97.29 | B.1.617.2 |
| 14941 | S3675-(ORF1a);G3676-(ORF1a);F3677-(ORF1a);H69-(S);V70-(S);Y144-(S) | 99.78 | B.1.1.7 |
| 14945 | D119-(ORF8);F120-(ORF8) | 69.39 | None |
| 14946 | D119-(ORF8);F120-(ORF8);F157-(S);R158-(S) | 99.18 | B.1.617.2 |
| 14947 | D119-(ORF8);F120-(ORF8);F157-(S);R158-(S) | 98.34 | B.1.404 |
| 14948 | D119-(ORF8);F120-(ORF8) | 60.82 | None |
| 14949 | D119-(ORF8);F120-(ORF8);F157-(S);R158-(S) | 99.83 | B.1.617.2 |
| 14950 | D119-(ORF8);F120-(ORF8);F157-(S);R158-(S) | 98.94 | B.1.617.2 |
| 14951 | D119-(ORF8);F120-(ORF8) | 36.15 | None |
| 14952 | D119-(ORF8);F120-(ORF8);F157-(S);R158-(S) | 98.97 | B.1.617.2 |
| 14953 | D119-(ORF8);F120-(ORF8);F157-(S);R158-(S) | 99.82 | B.1.617.2 |
| 14954 | D119-(ORF8);F120-(ORF8);F157-(S);R158-(S) | 99.45 | B.1.617.2 |
| 14955 | D119-(ORF8);F120-(ORF8);F157-(S);R158-(S) | 99.72 | B.1.617.2 |
| 14957 |  | 20.91 | None |
| 14958 | D119-(ORF8);F120-(ORF8);F157-(S);R158-(S) | 99.8 | B.1.617.2 |
| 14960 | D119-(ORF8);F120-(ORF8);F157-(S);R158-(S) | 99 | B.1.617.2 |
| 14963 | D119-(ORF8);F120-(ORF8) | 69.01 | None |
| 14965 | D119-(ORF8);F120-(ORF8);F157-(S);R158-(S) | 99.73 | B.1.617.2 |
| 14966 | D119-(ORF8);F120-(ORF8);F157-(S);R158-(S) | 99.61 | B.1.617.2 |
| 14967 | D119-(ORF8);F120-(ORF8) | 70.23 | None |
| 14968 | D119-(ORF8);F120-(ORF8) |  | None |
| 14969 | D119-(ORF8);F120-(ORF8);F157-(S);R158-(S) |  | B.1.617.2 |
| 14970 | D119-(ORF8);F120-(ORF8);F157-(S);R158-(S) | 98.53 | B.1.617.2 |
| 14971 | D119-(ORF8);F120-(ORF8);F157-(S);R158-(S) | 99.9 | B.1.617.2 |
| 14972 | C1732-(ORF1b);L1733-(ORF1b);C1734-(ORF1b) | 36.73 | None |
| 14973 | D119-(ORF8);F120-(ORF8);F157-(S);R158-(S) | 97.09 | B.1.617.2 |
| 14975 | D119-(ORF8);F120-(ORF8);F157-(S);R158-(S) | 99.55 | B.1.617.2 |
| 14978 | D119-(ORF8);F120-(ORF8);F157-(S);R158-(S) | 99.68 | B.1.617.2 |
| 14980 | D119-(ORF8);F120-(ORF8);F157-(S);R158-(S) | 99.82 | B.1.617.2 |
| 14999 | D119-(ORF8);F120-(ORF8);F157-(S);R158-(S) | 98.63 | B.1.617.2 |

**Table 4 –continued**
| **Sample** | **AA DELETIONS** | **coverage %** | **Pangolin Clade** |
| --- | --- | --- | --- |
| **15006** | D119-(ORF8);F120-(ORF8);F157-(S);R158-(S) | 97.86 | B.1.617.2 |
| **15012** | D119-(ORF8);F120-(ORF8);F157-(S);R158-(S) | 94.57 | B.1.617.2 |
| **15014** | D119-(ORF8);F120-(ORF8) |  | None |
| **15019** | L206-(M);D119-(ORF8);F120-(ORF8);F157-(S);R158-(S) |  | B.1.617.2 |
| **15024** | D119-(ORF8);F120-(ORF8);F157-(S);R158-(S) | 94.55 | B.1.617.2 |
| **15036** | D119-(ORF8);F120-(ORF8);F157-(S);R158-(S) | 97.86 | B.1.617.2 |
| **15043** | D119-(ORF8);F120-(ORF8) | 47.82 | None |
| **15046** | D119-(ORF8);F120-(ORF8);F157-(S);R158-(S) | 97.87 | B.1.617.2 |
| **15048** | H69-(S);V70-(S);Y144-(S) | 98.28 | B.1.1 |
| **15051** | D119-(ORF8);F120-(ORF8);F157-(S);R158-(S) | 98.38 | B.1.617.2 |
| **15053** | D119-(ORF8);F120-(ORF8);F157-(S);R158-(S) | 91.98 | B.1.617.2 |
| **15085** | D119-(ORF8);F120-(ORF8) | 33.42 |  |

### 3.5 Detection of the Alpha and Delta lineage in wastewater samples

The capability of the Alpha-Delta and confirmatory Orf8_119del_ assays to detect the presence of the Delta lineage RNA in sewage samples was evaluated using samples from different periods of the COVID19 pandemic, from December 2020 until mid-June 2021. As detailed in **Table 5**, the assay correctly identified the samples from 2020 as non-Alpha, non-Delta. Samples collected during the peak of the third morbidity wave in Israel were all identified as positive for the Alpha variant, and samples collected from July 2021 were all identified as positive for the Delta variant (**Table 5**). The Cq values of the N_D3L_ and S_157del_ reactions, when positive, were comparable to those of the E-sarbeco reaction, demonstrating sufficient sensitivity of these reactions with RNA extraction of wastewater. These results demonstrate the capability of the Alpha-Delta assay to detect the presence of SC-2 in wastewater and determine whether the Alpha or Delta lineages, or both, are circulating in the examined regions. In order to evaluate the capability of the ORF8_119del_ reaction to identify the Delta linage RNA in wastewater samples, it was applied to samples that were previously identified as non-Alpha, non-Delta, or as Delta positive. Samples selected randomly from 2020 were all negative for the presence of the ORF8_119del_ mutation, while recent samples from July 2021 were all identified as positive for the mutation (**Table 6**). Although the Cq values obtained with the ORF8_119del_ reaction were 2-3 cycles higher than those of the S_157del_ reaction in this test, all samples were correctly identified. Collectively, these results show that both the Alpha-Delta multiplex and the confirmatory ORF8_119del_ test can be used for examination of identification of the Alpha and Delta lineages in wastewater samples.

**Table 5.** Detection of Alpha and Delta lineage in wastewater samples using the Alpha-Delta assay. Samples collected during 2020 and during July 2021 were examined using the Alpha-Delta assay and the Orf8_119del_ confirmatory test. The Cq values for each reaction are shown. N/A – No Amplification.

| Sample | CVL E | N<sub>B.1.1.7</sub> | S-157del | Sample | CVL E | N<sub>B.1.1.7</sub> | S-157del |
| --- | --- | --- | --- | --- | --- | --- | --- |
| S-2299 | 35 | N/A | N/A | S-12689 | 28 | N/A | 29 |
| S-2302 | 32 | N/A | N/A | S-12694 | 28 | N/A | 29 |
| S-2307 | 32 | N/A | N/A | S-12698 | 30 | N/A | 31 |
| S-2310 | 30 | N/A | N/A | S-12733 | 28 | N/A | 29 |
| S-2314 | 31 | N/A | N/A | S-12737 | 26 | N/A | 27 |
| S-2318 | 30 | N/A | N/A | S-12741 | 28 | N/A | 28 |
| S-2328 | 31 | N/A | N/A | S-12745 | 29 | N/A | 30 |
| S-2332 | 31 | N/A | N/A | S-12754 | 27 | N/A | 28 |
| S-2336 | 31 | N/A | N/A | S-12762 | 30 | N/A | 30 |
| S-2340 | 32 | N/A | N/A | S-12766 | 29 | N/A | 29 |
| S-2344 | 30 | N/A | N/A | Env_128 | 31 | N/A | 29 |
| S-2349 | 33 | N/A | N/A | Env_129 | 33 | N/A | 31 |
| S-2353 | 32 | N/A | N/A | Env_130 | 31 | N/A | 30 |
| S-2357 | 32 | N/A | N/A | Env_134 | 33 | N/A | 30 |
| S-2382 | 30 | N/A | N/A | Env_138 | 32 | N/A | 29 |
| S-2386 | 31 | N/A | N/A | Env_140 | 30 | N/A | 28 |
| S-2390 | 30 | N/A | N/A | Env_143 | 31 | N/A | 30 |
| S-2394 | 34 | N/A | N/A | Env_144 | 29 | N/A | 28 |
| S-2399 | 31 | N/A | N/A | Env_145 | 30 | N/A | 29 |
| S-2402 | 32 | N/A | N/A | Env_146 | 36 | N/A | 33 |
| S-2407 | 31 | N/A | N/A | Env_152 | 35 | N/A | 30 |
| S-2424 | 30 | N/A | N/A | Env_163 | 29 | N/A | 30 |
| S-2457 | 32 | N/A | N/A | Env_168 | 32 | N/A | 29 |
| S-2461 | 34 | N/A | N/A | Env_172 | 33 | N/A | 32 |
| S-2480 | 32 | N/A | N/A | Env_173 | 30 | N/A | 31 |
| S-2493 | 33 | N/A | N/A | Env_177 | 30 | N/A | 32 |
| S-2498 | 30 | N/A | N/A | Env_186 | 30 | N/A | 31 |
| S-2501 | 29 | N/A | N/A | Env_197 | 29 | N/A | 29 |
| S-2505 | 31 | N/A | N/A | Env_198 | 30 | N/A | 30 |
| S-2509 | 33 | N/A | N/A | S-12686 | 31 | N/A | 31 |
| S-7369 | 31.50 | 32.74 | N/A |  |  |  |  |
| S-7419 | 31.36 | 33.01 | N/A |  |  |  |  |
| S-7451 | 31.79 | 33.40 | N/A |  |  |  |  |
| S-7455 | 31.01 | 33.16 | N/A |  |  |  |  |
| S-7459 | 30.63 | 31.79 | N/A |  |  |  |  |
| S-7463 | 30.83 | 32.03 | N/A |  |  |  |  |
| S-7467 | 31.26 | 32.39 | N/A |  |  |  |  |
| S-7592 | 31.21 | 32.19 | N/A |  |  |  |  |
| S-7600 | 31.11 | 32.39 | N/A |  |  |  |  |
| S-7604 | 31.76 | 33.44 | N/A |  |  |  |  |
| S-7613 | 32.14 | 33.61 | N/A |  |  |  |  |
| S-7678 | 32.75 | 34.74 | N/A |  |  |  |  |
| S-7699 | 29.06 | 30.15 | N/A |  |  |  |  |
| S-7718 | 32.71 | 34.16 | N/A |  |  |  |  |
| S-7722 | 31.89 | 33.72 | N/A |  |  |  |  |
| S-7727 | 31.78 | 33.10 | N/A |  |  |  |  |
| S-7734 | 32.72 | 33.49 | N/A |  |  |  |  |
| S-7742 | 32.71 | 33.92 | N/A |  |  |  |  |
| S-7754 | 32.02 | 33.40 | N/A |  |  |  |  |
| S-7762 | 32.07 | 33.69 | N/A |  |  |  |  |
| S-7369 | 31.50 | 32.74 | N/A |  |  |  |  |
| S-7419 | 31.36 | 33.01 | N/A |  |  |  |  |
| S-7451 | 31.79 | 33.40 | N/A |  |  |  |  |
| S-7455 | 31.01 | 33.16 | N/A |  |  |  |  |
| S-7459 | 30.63 | 31.79 | N/A |  |  |  |  |
| S-7463 | 30.83 | 32.03 | N/A |  |  |  |  |
| S-7467 | 31.26 | 32.39 | N/A |  |  |  |  |
| S-7592 | 31.21 | 32.19 | N/A |  |  |  |  |

**Table 6.** Detection of Delta lineage in wastewater samples using the Orf8_119del_ test. Randomly selected wastewater samples that were previously tested with the Alpha Delta assay, were subsequently examined using the confirmatory Orf8119del test. N/A – No Amplification.

| Sample | CVL E | S-157del | Orf8-119del |
| --- | --- | --- | --- |
| S-2480 | 32 | N/A | N/A |
| S-2493 | 33 | N/A | N/A |
| S-2498 | 30 | N/A | N/A |
| S-2501 | 29 | N/A | N/A |
| S-2505 | 31 | N/A | N/A |
| S-2509 | 33 | N/A | N/A |
| 12689 | 28 | 29 | 31 |
| 12694 | 28 | 29 | 31 |
| 12698 | 30 | 31 | 32 |
| 12733 | 28 | 29 | 31 |
| 12737 | 26 | 27 | 30 |
| 12741 | 28 | 28 | 31 |
| 12745 | 29 | 30 | 32 |
| 12754 | 27 | 28 | 30 |
| 12762 | 30 | 30 | 32 |
| 12766 | 29 | 29 | 32 |
| Env_128 | 31 | 29 | 32 |
| Env_129 | 33 | 31 | 34 |
| Env_130 | 31 | 30 | 32 |
| Env_134 | 33 | 30 | 34 |
| Env_138 | 32 | 29 | 31 |
| Env_140 | 30 | 28 | 31 |
| Env_143 | 31 | 30 | 33 |
| Env_144 | 29 | 28 | 30 |
| Env_145 | 30 | 29 | 31 |
| Env_146 | 36 | 33 | 33 |
| Env_152 | 35 | 30 | 33 |
| Env_163 | 29 | 30 | 32 |
| Env_168 | 32 | 29 | 32 |
| Env_172 | 33 | 32 | 35 |
| Env_173 | 30 | 31 | 34 |
| Env_177 | 30 | 32 | 34 |
| Env_186 | 30 | 31 | 34 |
| Env_197 | 29 | 29 | 32 |
| Env_198 | 30 | 30 | 33 |
| S-12686 | 31 | 31 | 34 |

## 4. DISCUSSION

The emergence of SC-2 variants that have the potential to affect both COVID-19 spreading and vaccine evasion, poses additional challenge for current intensive diagnostics efforts worldwide. The primary goal of diagnostic SC-2 tests was initially to detect the presence of SC-2 RNA, either in clinical or environmental samples. However, the need for rapid classification of SC-2 sample for their lineage, once a sample is determined as SC-2 positive, becomes essential, when attempting to monitor emergence or incursion of new variants, into a country, a geographic region, or a community. Surveillance through sequencing is probably the ultimate tool to identify the emergence of new and potentially important variants [19]. However, whole genome sequencing (WGS) is expensive, time-consuming, required exceptionally skilled personnel and cannot be readily scaled-up for high throughput testing, compared to rapid molecular tests [16,20]. In this dynamic situation of emerging variants, rapid, cost-effective and high throughput identification of circulating SC-2 variants is required.

We recently reported on the development and utilization of a rapid multiplex RT-qPCR assay that successfully classified SC-2 samples as Alpha or Beta, or neither. We demonstrated that it could be readily scaled-up to accommodate high throughput testing [17]. Here, we describe a new assay that combines general detection of SC-2, together with specific classification of the two major variants Alpha and Delta. Previous reports of in-house selective assays, as well as commercial variant detection kits, rely mostly on the identification of common mutations that are not specific to a single variant [21,22,23]. Other VOC detection approaches rely on multi-step expensive procedures [24,25]. Such assays are not suitable for rapid determination the identity of an examined sample, based on a single assay. Confirmation of the Alpha Delta assay results by Sanger and Illumina sequencing reinforced its importance as a first line rapid and reliable tool for classification of major VOCs, obviating the need for further examination. The identification of both Orf8 and Spike gene Delta-specific deletions, in samples with insufficient sequencing coverage, demonstrated the usefulness of the Alpha Delta assay, in determining the identity of “sequencing-resistant” samples. Such problems can result from low RNA quantity (as evident by the Cq values detailed in **Table 3**), or other factors preventing the completion of the sequencing procedure. This is particularly important in classification of wastewater-derived samples, where the target RNA is a pool from multiple individuals, is diluted and the sample contains chemical inhibitors. The Alpha Delta assay successfully detected the presence of the Delta variant even in very low RNA concentration, thereby enabling sensitive monitoring of the circulation of this variant during its incursion into the country. This assay was also used for screening international arrivals, which is particularly important when attempting to prevent the spreading of new variants. While other reported approaches to detect SC-2 variants in international travelers rely on lengthy multi-step procedures [24,25] this assay is rapid (∼80 minutes from extraction to answer) and can be implemented in most standard laboratories that perform routine PCR tests. This is particularly advantageous when an accelerated epidemiological investigation or other actions need to be taken, and complete genomic analysis is not available.

The Orf8_119del_ reaction was used in this study as a confirmatory assay, complementing the first-line Alpha Delta assay. This is useful to confirm ambiguous results, and to provide additional support for lineage classification, when sequencing analysis is not available. The S_157del_ and the Orf8_119del_ reactions showed similar performance in both clinical and wastewater samples and are therefore equally useful in the current situation where both deletions are unique for the Delta variant. However, if a new variant will emerge, where one of these mutations is present, re-assessment of the multiplex combination can be performed, in order to adjust the reactions to the required assay specificity. The ability to assemble different reactions into a single multiplex assay is advantageous with respect to the intended use of the assay, as we show here. When the Beta variant was introduced in Israel, we used the combination of the Alpha Beta and inclusive E reactions to classify the examined samples. Upon introduction of the Delta variant, we modified the assay and added a confirmatory reaction, to accommodate the pressing need to identify the Delta and Alpha variants, but also identify non-Alpha, non-Delta positive samples, all in a single 80-minutes standard RT-qPCR assay.

This study therefore demonstrates that in addition to implementation of a useful diagnostic tool, the approach of a developing tailor-made variant-specific qPCR assay can improve the SC-2 diagnostic capability in any qPCR-qualified laboratory, and can complement variant classification when WGS analysis either fails or is not available. Furthermore, such an assay can be valuable for epidemiological studies, enabling a quick and affordable assessment of the circulation of major variants in a community, or in a geographic region of interest. It is conceivable that the emergence of future SC-2 variants, and possibly other infectious viruses, will warrant further development of such diagnostic tools.

## Data Availability

All data produced in the present study are available upon reasonable request to the authors

## ETHICAL STATEMENT

The study was conducted according to the guidelines of the Declaration of Helsinki, and approved by the Institutional Review Board of the Sheba Medical Center institutional review board (7045-20-SMC). Patient consent was waived as the study used remains of clinical samples and the analysis used anonymous clinical data.

