## Supplementary material file for "SPECIFIC DETECTION OF SARS-COV-2 VARIANTS B.1.1.7 (ALPHA) AND B.1.617.2 (DELTA) USING A ONE-STEP QUANTITATIVE PCR ASSAY"

| Sample | COV19 E | $S_{157del}$ | $Orf8_{119del}$ |
|---|---|---|---|
| WT 1 | 27.36 | N/A | N/A |
| WT 2 | 31.1 | N/A | N/A |
| WT 3 | 25.27 | N/A | N/A |
| WT 4 | 24.99 | N/A | N/A |
| WT 5 | 28.04 | N/A | N/A |
| B.1.1.7 - 1 | 22.08 | N/A | N/A |
| B.1.1.7 - 2 | 18.73 | N/A | N/A |
| B.1.1.7 - 3 | 24.61 | N/A | N/A |
| B.1.351 - 1 | 19.3 | N/A | N/A |
| B.1.351 - 2 | 28.28 | N/A | N/A |
| B.1.351 - 3 | 20.34 | N/A | N/A |
| B.1.351 - 4 | 29.9 | N/A | N/A |
| P1 | 32.15 | N/A | N/A |

**Supplementary Table S1. Specificity evaluation of the B.1.617.2 $S_{157del}$ and the $Orf8_{119del}$ reactions.** Samples from indicated lineages were examined using a duplex assay consisting of the inclusive E-sarbeco reaction, and the specific $S_{156-157del}$ reaction and a different duplex assay containing the E-sarbeco and the $Orf8_{119del}$ reactions. The Cq values obtained for the E-sarbeco reaction are from the E+$S_{157de}$l test. N/A – No Amplification.

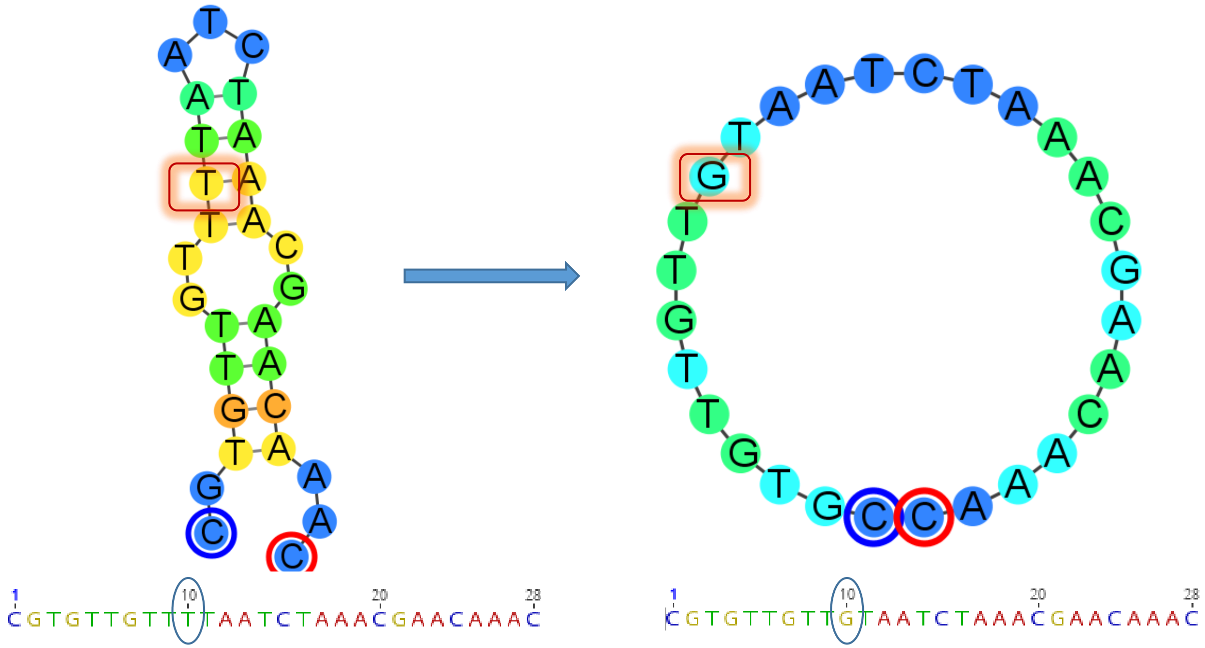


**<u>Supplementary Figure S1.</u> Design of the Orf8119-120del probe.** In order to avoid secondary structure formation of the 28199 probe, the deoxythymidine base was replaced with deoxyguanine, thereby destabilizing the secondary structure and enabling annealing of the probe to the target sequence. The base replacement positions are circled. The simulation temperature was 60°C. The simulation was performed using the Geneious software (www.geneious.com).

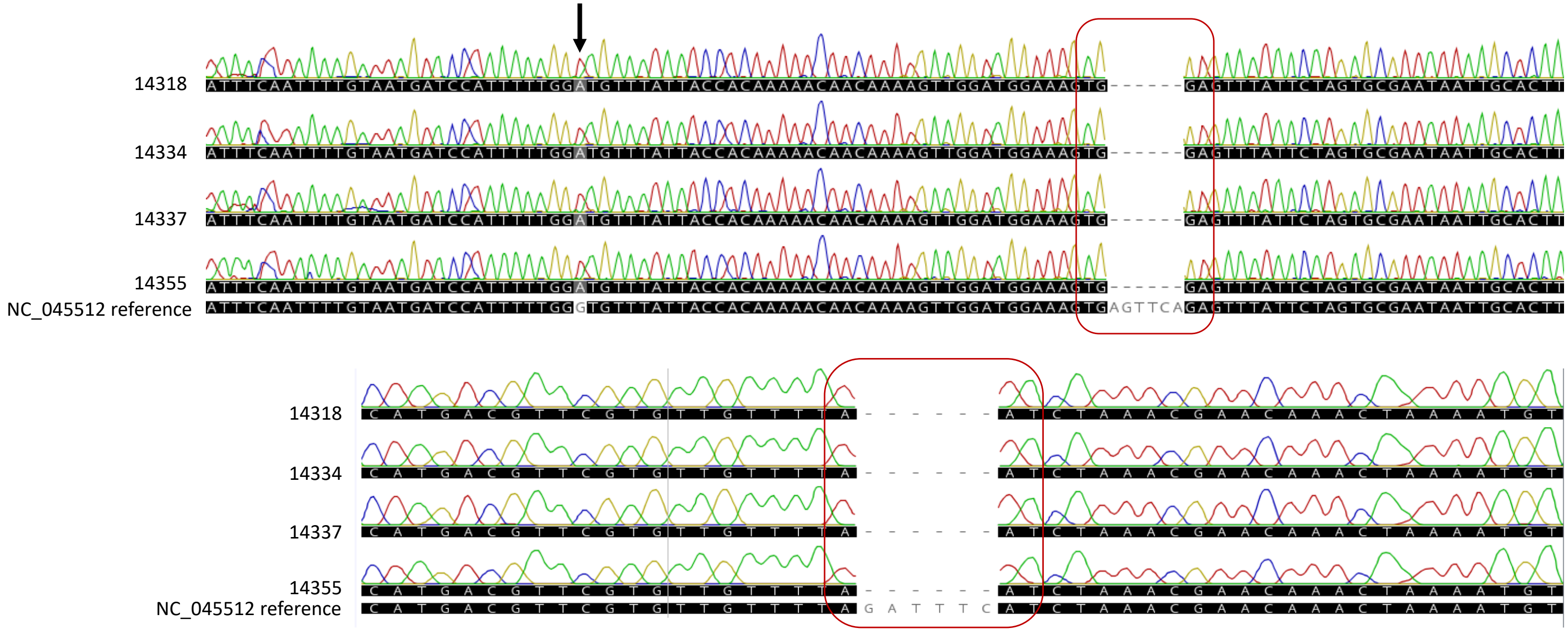


**<u>Supplementary Figure S2.</u> Sanger Sequencing of B.1.617-suspected samples.** The B.1.617-specific deletion regions of four samples identified as "B.1.617-suspected" by the $S_{157del}$ and $Orf8_{119del}$ reactions were sequenced. (A) Alignment of the $S_{157del}$ region with reference sequence NC_045512. The deletion region is marked with a rectangle. The A to G substitution, which translates into G142D amino acid mutation, is marked with an arrow. (B) Alignment of the $Orf8_{119del}$ region with reference sequence NC_045512. The deletion region is marked with a rectangle.

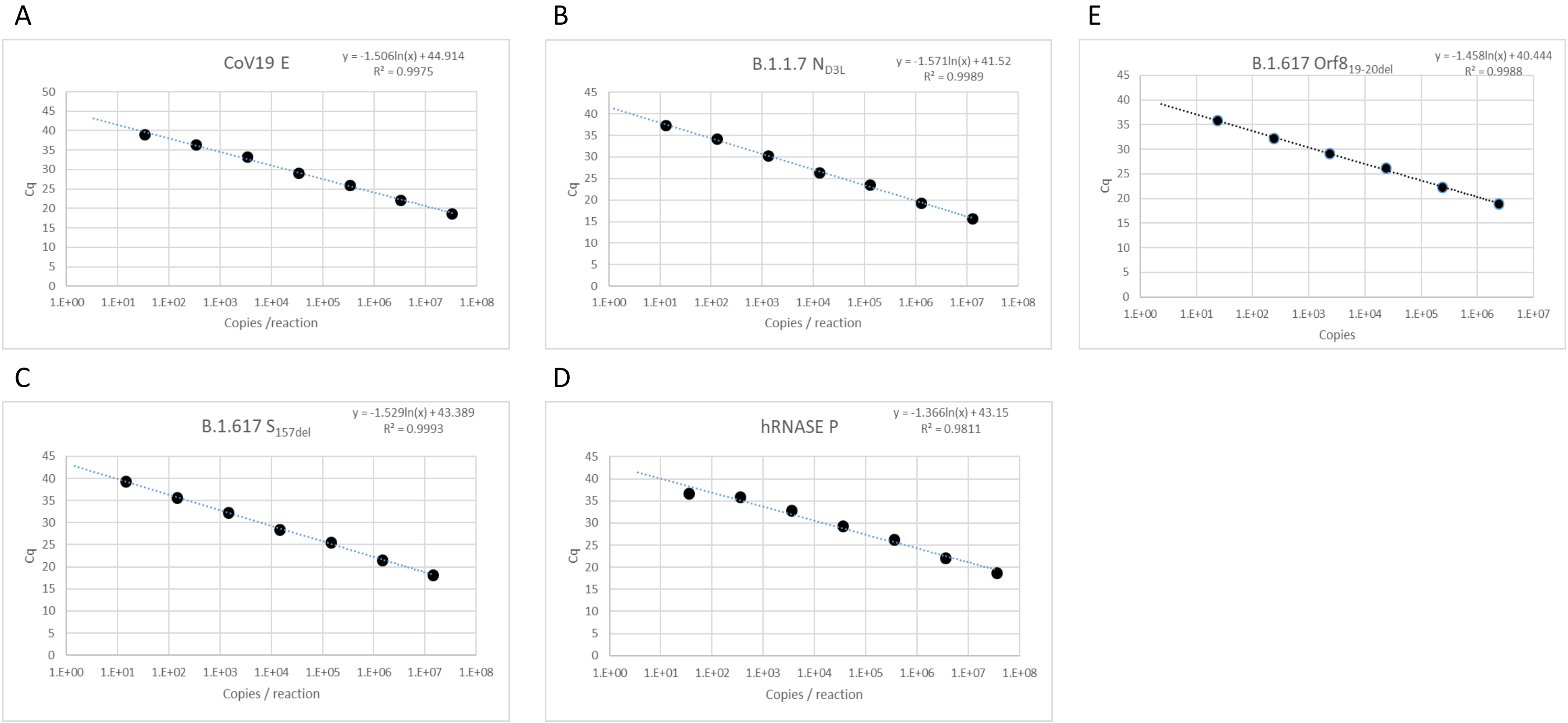


**<u>Supplementary Figure S3.</u>**Analytical Limit of Detection (LOD) of the Alpha-Delta assay**.** In vitro transcribed RNA targets of the four assay reactions were pooled and then diluted 10-fold. The average Cq value of each target for each dilution was plotted against the calculated target concentration. The derived regression curve, its formula and $R^2$ value are shown for each graph. (A) E-sarbeco reaction plot (B) $N_{D3L}$ reaction plot (C) $S_{157del}$ plot (D) hRNAse P plot. The samples were tested in triplicates.

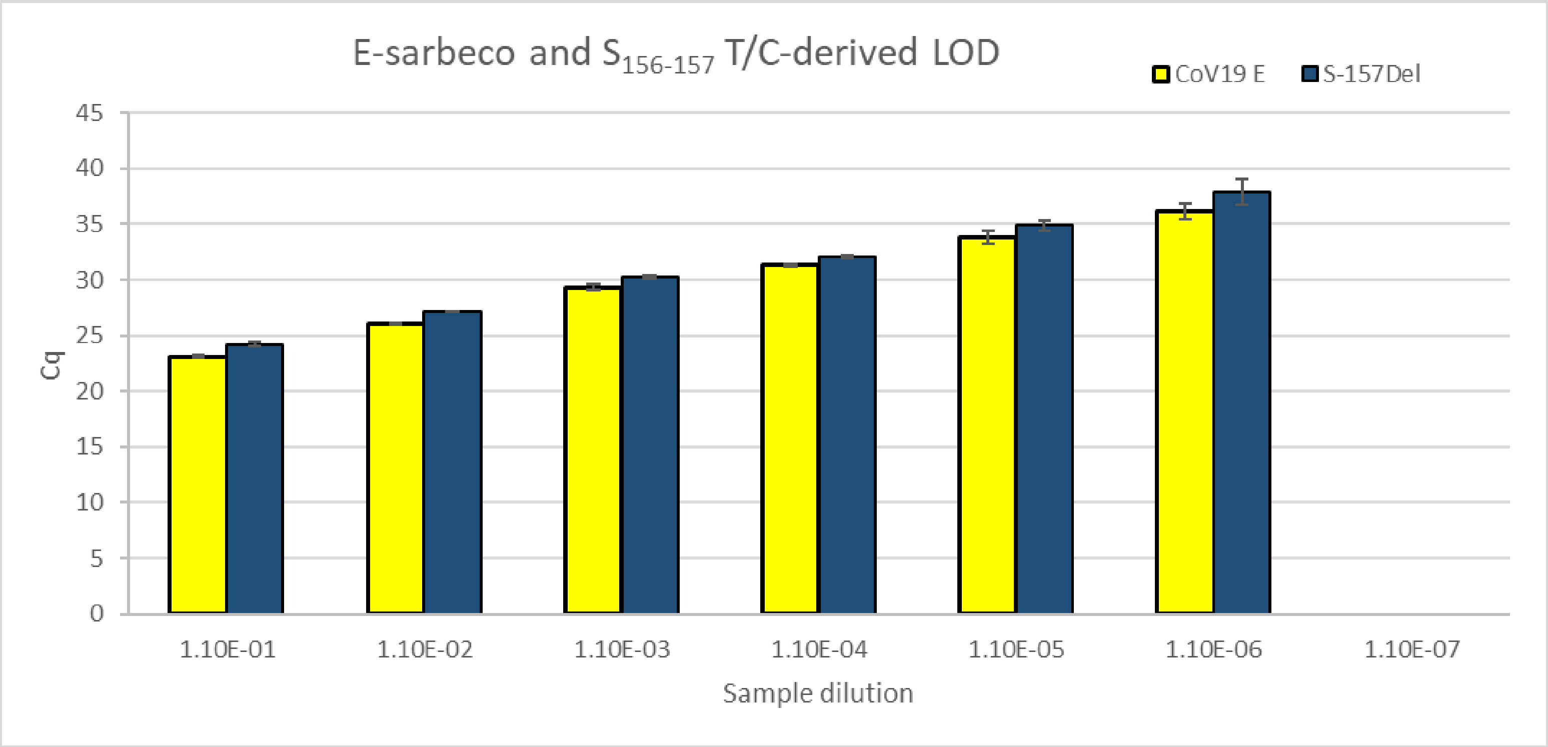


**<u>Supplementary Figure S4. Evaluation of the the</u> $S_{157del}$ reaction sensitivity with culture-derived virus.** RNA was extracted from cultured B.1.617.2 strain and serial dilutions were prepared as indicated. The dilutions were tested using the Alpha-Delta assay. The average Cq value and standard deviation of 3 repeats for each dilution are shown, for the inclusive E reaction and the specific $S_{157\text{-}158del}$ reaction.
